# Precision Oncology Insights into WNT Pathway Alterations in FOLFOX-Treated Early-Onset Colorectal Cancer in High-Risk Populations

**DOI:** 10.1101/2025.08.02.25332877

**Authors:** Fernando C. Diaz, Brigette Waldrup, Francisco G. Carranza, Sophia Manjarrez, Enrique Velazquez-Villarreal

## Abstract

**Background:** Early-onset colorectal cancer (EOCRC), defined as diagnosis before age 50, is rising rapidly and disproportionately affects high-risk populations, including Hispanic/Latino (H/L) individuals. Although FOLFOX is a standard first-line chemotherapy for microsatellite stable (MSS) CRC, its efficacy and impact on tumor genomics in EOCRC remain understudied. Given the central role of WNT signaling in CRC pathogenesis, this study aimed to characterize WNT pathway alterations in EOCRC across diverse populations and assess their association with FOLFOX treatment and clinical outcomes.

**Methods:** We analyzed somatic mutation data from 2,515 CRC patients (266 H/L, 2,249 Non-Hispanic White [NHW]) from publicly available datasets. Patients were stratified by age (EOCRC vs. late-onset CRC [LOCRC]), ancestry (H/L vs. NHW), and FOLFOX treatment status. Mutation frequencies in WNT pathway genes were compared using chi-squared tests, and Kaplan-Meier analysis was used to assess overall survival.

**Results:** WNT pathway alterations were prevalent across all groups, with APC mutations dominating. Among H/L EOCRC patients, FOLFOX-treated individuals had significantly lower mutation rates in CTNNB1 (5.5% vs. 17.3%, p = 0.04) and RNF43 (5.5% vs. 19.2%, p = 0.02) compared to untreated counterparts. Similar patterns were observed between untreated EOCRC and LOCRC H/L patients for CTNNB1 (17.3% vs. 4.0%, p = 0.05) and RNF43 (19.2% vs. 10.0%, p = 0.006). In NHW LOCRC patients, FOLFOX treatment was associated with lower mutation frequencies in AXIN1 (2.0% vs. 4.6%, p = 0.004), AXIN2 (3.5% vs. 9.8%, p = 4.4×10⁻⁷), RNF43 (6.5% vs. 14.7%, p = 1.5×10⁻⁷), and TCF7L2 (13.1% vs. 18.1%, p = 0.008). Among NHW EOCRC patients, APC mutations were more frequent in untreated vs. treated groups (81.1% vs. 73.2%, p = 0.01), while AXIN2 and RNF43 mutations were less common. H/L EOCRC patients not treated with FOLFOX had significantly higher CTNNB1 (17.3% vs. 7.6%, p = 0.047) and RNF43 (19.2% vs. 6.6%, p = 0.006) mutation frequencies than their NHW counterparts. Kaplan-Meier analysis showed that WNT pathway alterations were associated with improved overall survival in FOLFOX-treated NHW EOCRC patients (p = 0.025) and in both treated (p = 0.02) and untreated (p = 0.015) NHW LOCRC patients. No significant survival differences were observed in H/L patients, though favorable trends were noted.

**Conclusions:** WNT pathway dysregulation is pervasive in EOCRC and varies by ancestry, treatment status, and age. FOLFOX chemotherapy appears to reduce the prevalence of specific non-canonical WNT alterations (e.g., CTNNB1, RNF43) in H/L EOCRC, while in NHW patients, broader WNT gene reductions were associated with improved survival. These findings suggest ancestry-specific molecular responses to chemotherapy and underscore the need for precision oncology approaches tailored to high-risk populations.

## 1. Introduction

Colorectal cancer (CRC) is the third most commonly diagnosed cancer in the United States and remains a leading cause of cancer-related deaths, ranking third among men and fourth among women [1]. While overall CRC incidence has declined in high-income nations due to improved screening programs [2], early-onset colorectal cancer (EOCRC)—defined as CRC diagnosed before the age of 50—has seen a sharp and concerning increase [3–5]. Alarmingly, projections estimate that EOCRC will become the leading cause of cancer-related death in individuals aged 20–49 by 2030 [4]. Stratified analyses reveal that EOCRC incidence is rising across all racial and ethnic groups, with particularly significant increases observed among Hispanic/Latino (H/L) [6–9], non-Hispanic Black, and American Indian/Alaska Native populations [10–12]. These trends reflect broader disparities in cancer care and outcomes, warranting dedicated investigation into the molecular features of EOCRC within high-risk and underserved populations.

Emerging studies suggest that EOCRC is biologically distinct from late-onset CRC (LOCRC), exhibiting divergent genetic and clinical features. Some reports have identified elevated tumor mutation burden, microsatellite instability (MSI), and increased PD-L1 expression in EOCRC, while others have challenged the consistency of these findings [13–15]. Nonetheless, comparative analyses indicate significant differences in the mutational profiles of genes involved in key oncogenic pathways, including TP53, SMAD4, BRAF, NOTCH1, CTNNB1, APC, and KRAS [13, 14, 16]. Additionally, LINE-1 hypomethylation has been proposed as a unique molecular marker of EOCRC [17]. These inconsistencies in the literature underscore the need for further research to define the molecular landscape of EOCRC and its clinical relevance—particularly in high-risk populations where genomic data remain sparse.

EOCRC is often diagnosed at more advanced stages and associated with poorer outcomes than LOCRC, with a higher probability of progression to metastatic disease [18, 19]. The current standard-of-care treatment for metastatic, microsatellite stable (MSS), and proficient mismatch repair (pMMR) CRC lacking targetable mutations is FOLFOX, a combination of folinic acid, 5-fluorouracil (5-FU), and oxaliplatin [20, 21]. However, recent data suggest that patients with EOCRC may experience more severe toxicity and shorter overall survival when treated with FOLFOX compared to those with LOCRC [22]. Despite these findings, little is known about the specific genomic drivers that might underlie these differential responses to therapy, especially in MSS EOCRC.

One pathway critically involved in CRC progression and chemoresistance is the WNT signaling pathway [23, 24]. The WNT pathway regulates β-catenin activation and is commonly disrupted through mutations in genes such as APC, CTNNB1, and RNF43 [25]. Over 80% of CRC tumors harbor mutations in APC, highlighting its central role in WNT dysregulation [25]. In EOCRC, particularly among MSS tumors, recent evidence suggests a lower prevalence of WNT pathway mutations relative to LOCRC [16], but paradoxically, higher β-catenin activation has been observed [26]. The functional consequences of this activation, especially in the context of standard chemotherapy, remain unclear. Preclinical models have shown minimal to moderate modulation of WNT signaling following FOLFOX treatment in tumors with elevated pathway activity [27]. However, these observations require clinical validation to determine their translational relevance.

Given the rising burden of EOCRC among high-risk populations such as H/L individuals and the limited understanding of how WNT pathway alterations affect treatment outcomes, this study seeks to characterize the molecular landscape of WNT signaling in MSS EOCRC treated with FOLFOX. By integrating genomic profiling with clinical outcome data, we aim to identify pathway-specific alterations that may influence treatment response and inform precision oncology strategies tailored to underserved populations. Understanding the distinct molecular features of MSS EOCRC, particularly within the context of WNT dysregulation, holds promise for guiding therapeutic interventions and addressing disparities in CRC care.

## 2. Materials and Methods

### 2.1 Clinical and genomic data

This study utilized clinical and genomic data from three publicly available CRC datasets curated through the cBioPortal for Cancer Genomics. To ensure the inclusion of detailed treatment information, datasets were selected based on the availability of chemotherapy and clinical annotation fields. The datasets included: Colorectal Adenocarcinoma (TCGA, PanCancer Atlas), MSK-CHORD (MSK, Nature 2024), and GENIE BPC CRC. Each dataset encompassed cases of colorectal, colon, and rectal adenocarcinomas. To maintain consistency, we restricted analysis to primary tumor samples and allowed only one tumor sample per patient.

Patients were identified as Hispanic/Latino (H/L) based on annotations such as “Hispanic or Latino,” “Spanish, NOS,” “Hispanic, NOS,” or “Latino, NOS.” Individuals with Mexican or Spanish surnames were also included following validated surname-based classification methods. The resulting cohort comprised 266 H/L patients (125 EOCRC and 141 LOCRC) and 2,249 Non-Hispanic White (NHW) patients (677 EOCRC and 1,572 LOCRC), all meeting identical inclusion criteria (Tables 1 & 2). Age at diagnosis was extracted from clinical metadata. EOCRC was defined as diagnosis before age 50, while LOCRC was defined as diagnosis at age 50 or older.

**Table 1.**
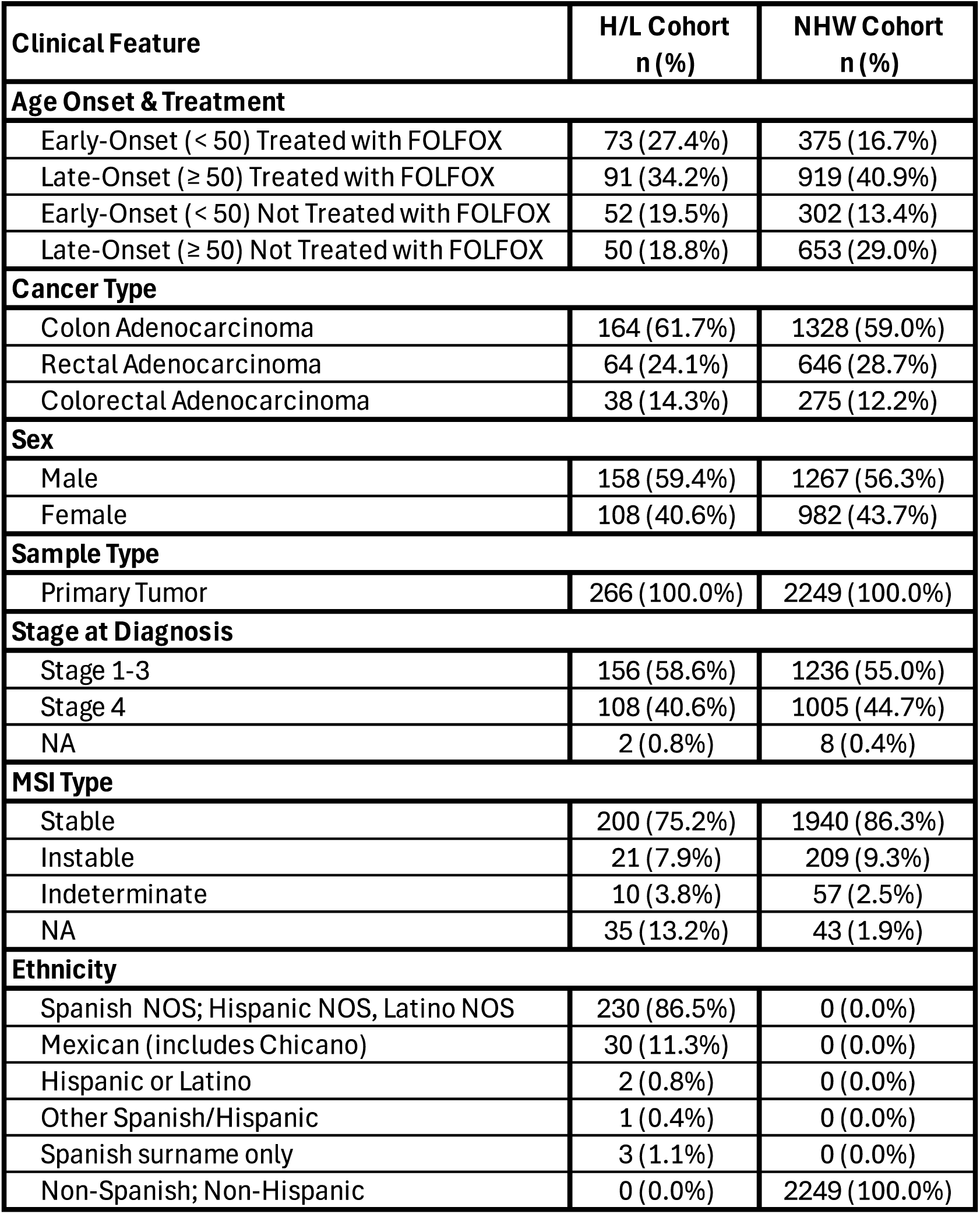
Clinical and demographic profiles of Hispanic/Latino (H/L) and Non-Hispanic White (NHW) colorectal cancer (CRC) patients in relation to age at onset, FOLFOX treatment, tumor characteristics, and ethnicity.

**Table 2.**
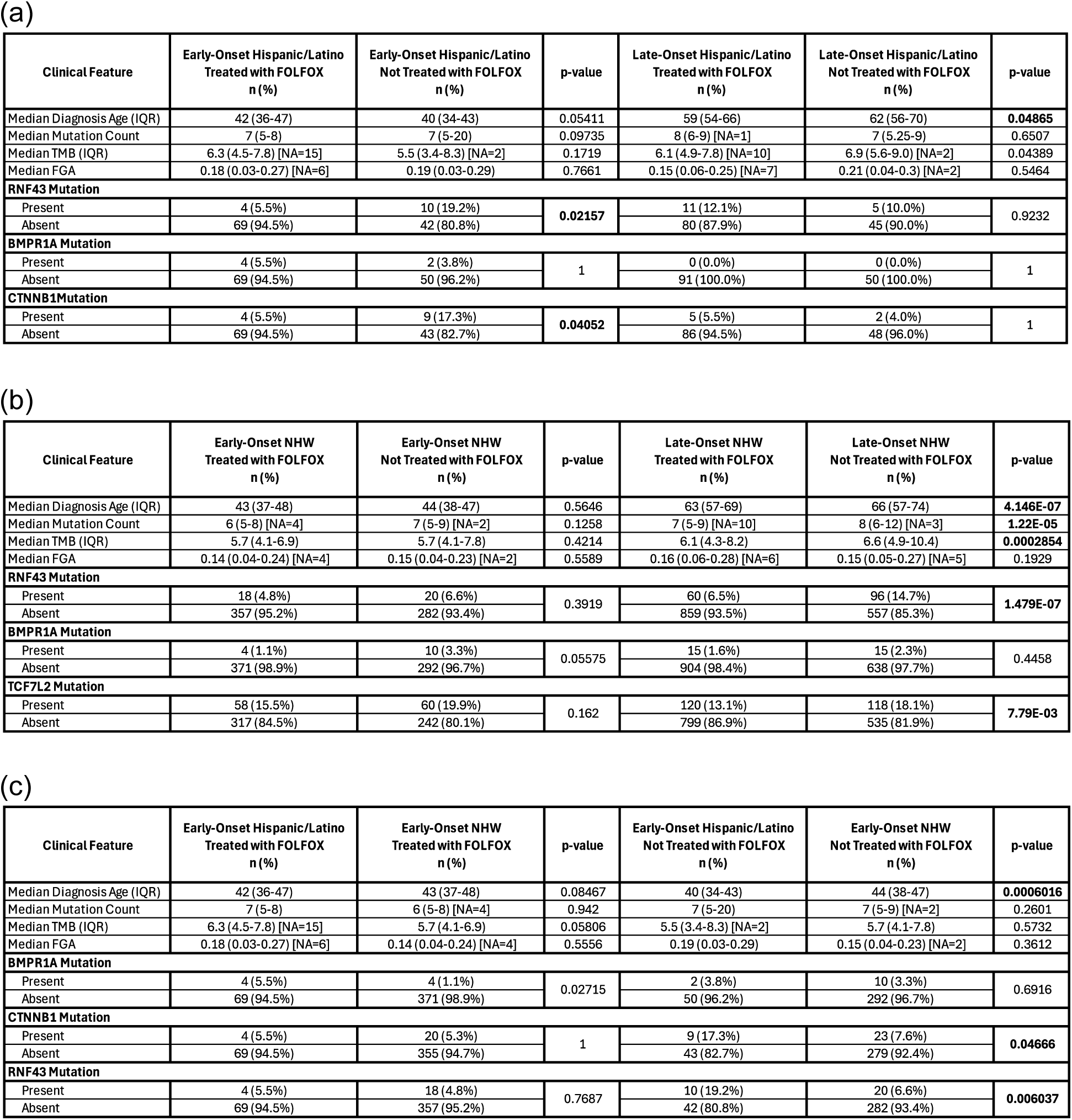
Comparative clinical and genomic characteristics across early-onset and late-onset colorectal cancer (CRC) patient cohorts. This table summarizes clinical and molecular differences in WNT pathway alterations and mutation burden across key subgroups: (a) Early-Onset Colorectal Cancer (EOCRC) vs. Late-Onset Colorectal Cancer (LOCRC) among Hispanic/Latino (H/L) patients; (b) EOCRC vs. LOCRC among Non-Hispanic White (NHW) patients; and (c) EOCRC comparisons between H/L and NHW patient cohorts. Each comparison highlights age at diagnosis, total mutation count, and frequencies of select WNT pathway gene alterations, stratified by ethnicity and age group.

To evaluate the role of treatment, cases were classified as “FOLFOX-treated” if they received a combination of leucovorin, fluorouracil (5-FU), and oxaliplatin administered within overlapping treatment timelines. Treatment data were manually curated from clinical drug administration tables provided in each dataset. Patients were further stratified by treatment status (FOLFOX-treated vs. non-treated), ethnicity (H/L vs. NHW), age group (EOCRC vs. LOCRC), and presence or absence of WNT pathway alterations.

Molecular alterations in the WNT pathway were defined using curated gene lists from established CRC literature [6,7]. Genes with well-characterized roles in WNT signaling and colorectal tumorigenesis were included. Somatic alterations were extracted from cBioPortal and limited to nonsynonymous variants, including missense, nonsense, frameshift insertions or deletions, splice site mutations, and translation start site mutations. Pathway alteration burden was defined as the presence of at least one qualifying mutation in any WNT pathway member gene. Table 3 outlines the distribution of WNT pathway alterations by age, ethnicity, and treatment group. Table 4 further compares EOCRC cases between H/L and NHW patients stratified by treatment type, enabling an in-depth comparative analysis.

**Table 3.**
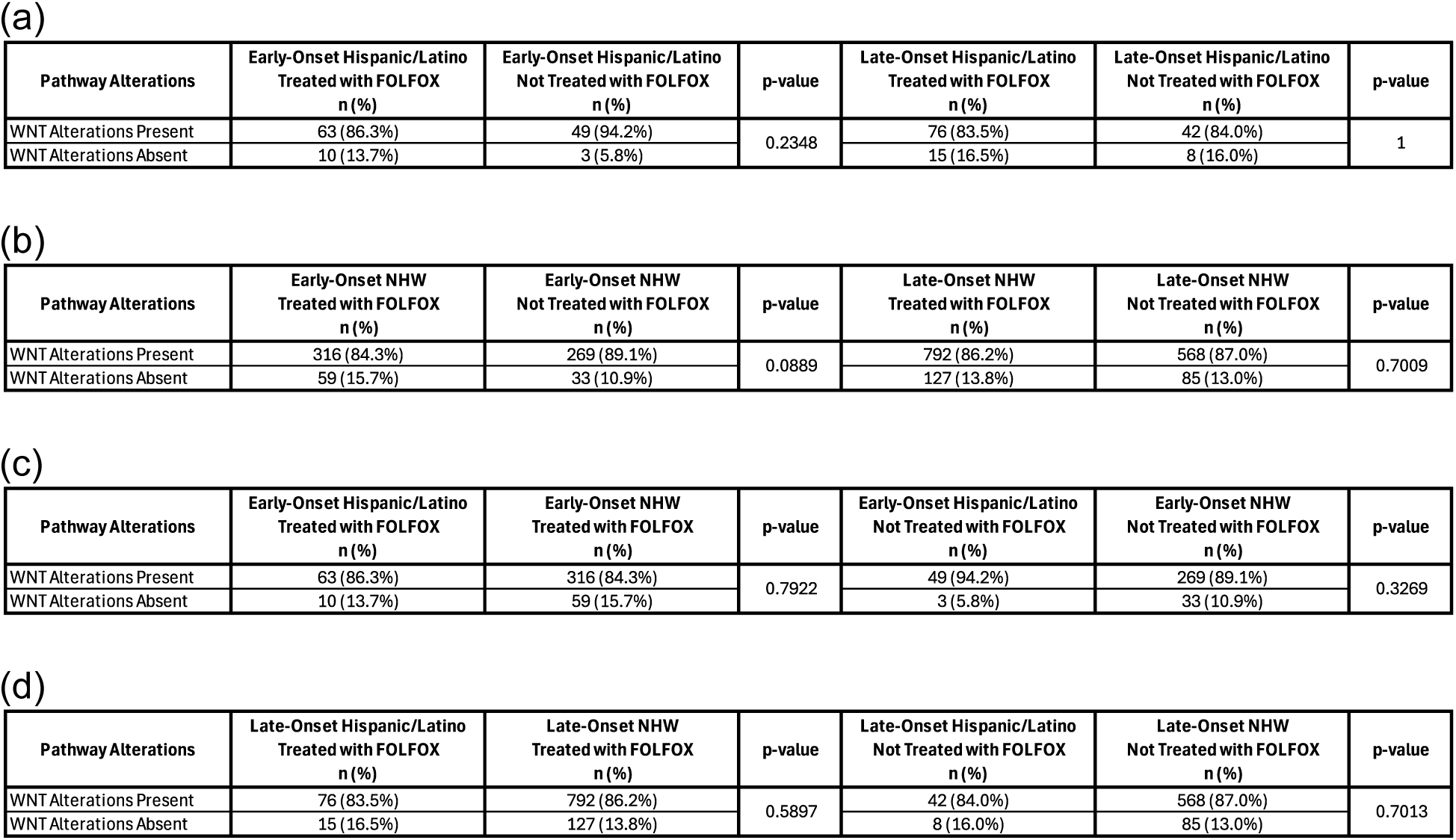
Frequency of WNT pathway alterations in Hispanic/Latino colorectal cancer (CRC) patients stratified by age of onset and FOLFOX treatment status. This table summarizes the mutation frequencies of key genes involved in the WNT signaling pathway among Hispanic/Latino (H/L) CRC patients, stratified by (3a) early-onset (EOCRC) vs. late-onset (LOCRC) and FOLFOX treatment status within the H/L cohort, (3b) FOLFOX-treated vs. untreated patients within EOCRC and LOCRC subgroups, (3c) EOCRC H/L vs. Non-Hispanic White (NHW) patients by FOLFOX treatment status, and (3d) LOCRC H/L vs. NHW patients by FOLFOX treatment status. Genes analyzed include *APC*, *CTNNB1*, *RNF43*, *AXIN1*, *AXIN2*, *AMER1*, and *TCF7L2*. Statistically significant differences (*p* < 0.05, Chi-square or Fisher’s exact test) are indicated with asterisks. This stratified analysis highlights potential interactions between age, ancestry, chemotherapy exposure, and WNT pathway dysregulation.

### 2.2 Statistical analysis

To assess differences in mutation prevalence across demographic and treatment groups, chi-square (χ²) tests were conducted to evaluate associations between categorical variables, including age group, ethnicity, FOLFOX treatment status, and WNT pathway alteration status. When expected cell counts were <5, Fisher’s exact tests were employed to ensure robust statistical inference. Mutation rates for individual WNT pathway genes were calculated for each subgroup, and gene-by-gene comparisons were performed to identify significant differences in mutation frequency.

Tumor samples were also categorized by anatomical site (colon vs. rectal adenocarcinoma) to explore potential interactions between tumor location, WNT pathway status, and treatment response. Supplementary Tables S1 and S2 provide detailed gene-level mutation frequencies by age, ethnicity, and treatment type, supporting interpretation of molecular trends within and across subgroups.

Kaplan–Meier survival analyses were performed to determine the impact of WNT pathway alterations on overall survival in EOCRC and LOCRC patients, stratified by treatment and ethnicity. Survival curves were generated to visualize differences in survival probability over time between groups with and without WNT alterations. The log-rank test was used to assess statistical significance between survival distributions.

Median survival times and 95% confidence intervals (CIs) were calculated to ensure accurate estimates. This multi-layered analytical framework provides a comprehensive evaluation of WNT pathway dysregulation in CRC, particularly in the context of FOLFOX treatment and underrepresented populations.

## 3. Results

### 3.1 Clinical and demographic characteristics of Hispanic/Latino and Non-Hispanic White CRC Cohorts

Using data from three cBioPortal projects with ethnicity and treatment annotations, we identified two study cohorts: 266 Hispanic/Latino (H/L) patients and 2,249 Non-Hispanic White (NHW) patients with primary colorectal, colon, or rectal adenocarcinoma. Within the H/L cohort, 27.4% (n = 73) were EOCRC patients treated with FOLFOX, 34.2% (n = 91) were LOCRC patients treated with FOLFOX, 19.5% (n = 52) were EOCRC without FOLFOX treatment, and 18.8% (n = 50) were LOCRC without FOLFOX treatment. In the NHW cohort, a smaller proportion (16.7%, n = 375) were EOCRC patients treated with FOLFOX, while 40.9% (n = 919) were LOCRC treated with FOLFOX. The remainder included 13.4% (n = 302) EOCRC and 29.0% (n = 653) LOCRC patients not treated with FOLFOX.

The sex distribution was similar across groups, with males comprising 59.4% of the H/L cohort and 56.3% of the NHW cohort. Females accounted for 40.6% of H/L patients and 43.7% of NHW patients. All patients included in this analysis had primary tumors, with 100% of both H/L and NHW patients meeting this criterion.

Regarding cancer type, most patients were diagnosed with colon adenocarcinoma— 61.7% in the H/L group and 59.0% in the NHW group. Rectal adenocarcinoma accounted for 24.1% of H/L cases and 28.7% of NHW cases, while colorectal adenocarcinoma (not otherwise specified) comprised 14.3% and 12.2%, respectively.

Stage at diagnosis varied between groups. Among H/L patients, 58.6% were diagnosed with stage 1–3 disease, while 40.6% were diagnosed at stage 4. A small proportion (0.8%) had missing or unreported stage data. In the NHW cohort, 55.0% of patients had stage 1–3 disease, 44.7% had stage 4, and only 0.4% lacked stage information.

Ethnicity sub-classification within the H/L cohort showed that the majority (86.5%) were labeled as Spanish NOS, Hispanic NOS, or Latino NOS. An additional 11.3% were identified as Mexican (including Chicano), and 2.2% were classified as Other Spanish/Hispanic. In contrast, all 2,249 NHW patients (100%) were confirmed as non-Hispanic White, ensuring clear separation between cohorts for comparative genomic and treatment analyses.

These clinical and demographic characteristics underscore distinct patterns of early-onset disease, treatment exposure, and diagnostic stage between H/L and NHW patients. The higher proportion of EOCRC among H/L patients, coupled with variation in cancer type and staging, reinforces the importance of stratified molecular analysis to inform precision oncology approaches in underserved populations.

### 3.2 Comparative genomic analysis by age and ancestry

A comparative analysis of clinical and genomic characteristics revealed key differences in early-onset (EOCRC) versus late-onset colorectal cancer (LOCRC) among Hispanic/Latino (H/L) patients (Table 2a). EOCRC H/L patients had a significantly younger median age at diagnosis (42 years; IQR: 37–46) compared to LOCRC H/L patients (66 years; IQR: 58–73, *p* < 0.001). The median mutation burden was slightly lower in EOCRC (7 mutations) than in LOCRC (8 mutations), though not statistically significant (*p* = 0.21). Notably, CTNNB1 mutations were significantly more frequent in EOCRC H/L patients (9.6%) than in LOCRC H/L patients (2.1%, *p* < 0.05). APC mutations were more common in LOCRC (73.8%) than in EOCRC (62.4%), but this difference did not reach significance (*p* = 0.06). RNF43 mutations trended higher in EOCRC (12.0%) compared to LOCRC (8.5%), though the difference was not statistically significant (*p*= 0.33).

Among Non-Hispanic White (NHW) patients (Table 2b), similar age-related patterns emerged. The median age at diagnosis for EOCRC was 42 years (IQR: 37–46), compared to 67 years (IQR: 59–75) for LOCRC (*p* < 0.001). Mutation burden was again slightly lower in EOCRC (median = 7) versus LOCRC (median = 8), without statistical significance (*p*= 0.18). APC mutations were significantly less frequent in EOCRC (67.1%) than in LOCRC (75.9%, *p* < 0.01). Conversely, CTNNB1 mutations were more common in EOCRC (7.4%) than LOCRC (2.8%, *p* < 0.01). RNF43 mutations were modestly enriched in EOCRC (6.7%) compared to LOCRC (4.3%), though this difference was not statistically significant (*p* = 0.08).

When comparing EOCRC patients across ancestry groups (Table 2c), the median age at diagnosis was identical (42 years; IQR: 37–46, *p* = 0.89), and mutation burdens were comparable (median = 7 in both groups, *p* = 0.98). However, RNF43 mutations were significantly more frequent in EOCRC H/L patients than in NHW patients (12.3% vs. 6.7%, *p* < 0.05), suggesting a potential ancestry-specific enrichment. CTNNB1 mutations were also higher in H/L (9.6%) than NHW (7.4%) patients, though the difference was not significant (*p* = 0.43). APC mutations were slightly less prevalent in EOCRC H/L patients (62.4%) compared to NHW patients (67.1%, *p* = 0.33).

### 3.3 Prevalence of WNT pathway alterations by age, ancestry, and FOLFOX treatment status

An integrated analysis of WNT pathway alterations revealed a consistently high prevalence across all subgroups of CRC patients, regardless of age of onset, ancestry, or FOLFOX treatment status.

In the Hispanic/Latino (H/L) cohort (Table 3a), WNT pathway alterations were present in 86.3% of EOCRC patients treated with FOLFOX and in 94.2% of untreated EOCRC cases, though this difference was not statistically significant (*p* = 0.23). Among late-onset CRC (LOCRC) patients, 83.5% of FOLFOX-treated and 84.0% of untreated individuals harbored WNT alterations (*p* = 1.00). The proportion of patients without WNT alterations was slightly higher in treated EOCRC (13.7%) compared to untreated (5.8%), and nearly identical in LOCRC (16.5% vs. 16.0%). These findings indicate that WNT pathway dysregulation is widespread in H/L CRC, with minimal variation by age or chemotherapy exposure.

Similarly, in the Non-Hispanic White (NHW) cohort (Table 3b), WNT alterations were detected in 84.3% of FOLFOX-treated EOCRC patients and 89.1% of untreated cases (*p* = 0.0889). Among LOCRC patients, prevalence was nearly identical between treated (86.2%) and untreated (87.0%) groups (*p* = 0.7009). Across all NHW subgroups, WNT alterations remained consistently high, and the proportion of patients without such alterations ranged from 10.9% to 15.7%, suggesting limited impact of FOLFOX on overall WNT mutational burden.

When comparing ancestry groups directly within the EOCRC population (Table 3c), the frequency of WNT alterations remained comparable between H/L and NHW patients. Among those treated with FOLFOX, 86.3% of H/L and 84.3% of NHW EOCRC patients had WNT pathway alterations (*p* = 0.7922). In untreated patients, 94.2% of H/L and 89.1% of NHW patients harbored such alterations (*p* = 0.3269). The proportion of patients without WNT alterations was low and similar across both groups.

Likewise, in the LOCRC subgroup (Table 3d), no significant differences were observed between H/L and NHW patients. WNT alterations were found in 83.5% of FOLFOX-treated H/L patients and 86.2% of their NHW counterparts (*p* = 0.5897), while the untreated group showed 84.0% and 87.0% prevalence, respectively (*p* = 0.7013). Together, these findings demonstrate that WNT pathway alterations are highly prevalent across early– and late-onset CRC, independent of ancestry or chemotherapy exposure. While specific gene-level differences may exist, the overall burden of WNT dysregulation appears to be a common and persistent feature of CRC biology in both Hispanic/Latino and Non-Hispanic White populations.

### 3.4 Frequencies of Gene Alterations in the WNT Pathway

#### Gene-level WNT pathway alterations in early-onset Hispanic/Latino patients by FOLFOX treatment

To assess treatment-associated genomic differences, WNT pathway mutation frequencies were evaluated in EOCRC patients of Hispanic/Latino (H/L) ancestry, stratified by FOLFOX exposure (Table S1). Notably, mutations in CTNNB1 and RNF43 were significantly more frequent in untreated patients (17.3% and 19.2%, respectively) compared to FOLFOX-treated patients (5.5% for both; p = 0.0405 and p = 0.0216). These findings suggest a potential selective effect of chemotherapy on non-canonical WNT components. In contrast, APC mutations were consistently high in both treated (80.8%) and untreated (84.6%) patients (p = 0.756), confirming its central role. Other genes (AMER1, AXIN1, AXIN2, GSK3B, TCF7L2) showed low mutation frequencies with no significant differences by treatment.

#### Gene-Level WNT pathway alterations in late-onset Hispanic/Latino patients by FOLFOX treatment

Among LOCRC H/L patients (Table S2), no statistically significant differences in WNT gene alterations were observed between FOLFOX-treated and untreated groups. APC mutations were the most frequent in both cohorts (71.4% vs. 72.0%; p = 1.00). RNF43 and AMER1 mutations showed slightly higher frequencies in treated patients (12.1% and 11.0%) compared to untreated patients (10.0% and 4.0%), though not statistically significant. Overall, FOLFOX did not appear to meaningfully impact the WNT mutation landscape in LOCRC H/L patients.

#### Comparison between early– and late-onset Hispanic/Latino patients treated with FOLFOX

In comparing WNT gene alterations between EOCRC and LOCRC H/L patients who received FOLFOX (Table S3), no significant differences were found. APC mutations were again the most prevalent (80.8% in EOCRC vs. 71.4% in LOCRC; p = 0.2266). Although RNF43 and AMER1 mutations were more common in LOCRC, these differences were not statistically significant. The distribution of CTNNB1, AXIN2, TCF7L2, and GSK3B mutations was similar across both age groups.

#### Comparison between untreated early– and late-onset Hispanic/Latino patients

Among untreated H/L patients (Table S4), RNF43 mutations were significantly more frequent in EOCRC than in LOCRC (19.2% vs. 10.0%; p = 0.0066). CTNNB1 mutations were also higher in EOCRC (17.3% vs. 4.0%), approaching significance (p = 0.0519). These findings point to a potential enrichment of non-canonical WNT pathway activity in younger patients. APC mutations were common across both groups. Other genes did not show statistically significant age-related differences.

#### Gene-level WNT alterations in early-onset Non-Hispanic White patients by FOLFOX treatment

In early-onset NHW patients (Table S5), no significant differences in mutation frequencies were found between FOLFOX-treated and untreated individuals. APC mutations were the most frequent (77.6% vs. 81.1%; p = 0.304). CTNNB1, RNF43, and TCF7L2 mutations were observed at moderate levels with no significant differences. Low-frequency mutations in AMER1, AXIN1, AXIN2, and GSK3B were consistent across treatment groups.

#### Gene-level WNT alterations in late-onset Non-Hispanic White patients by FOLFOX treatment

In LOCRC NHW patients (Table S6), FOLFOX treatment was associated with significantly lower mutation frequencies in several non-canonical WNT genes. AXIN1 (2.0% vs. 4.6%; p = 0.0045), AXIN2 (3.5% vs. 9.8%; p = 4.44×10⁻⁷), RNF43 (6.5% vs. 14.7%; p = 1.48×10⁻⁷), and TCF7L2 (13.1% vs. 18.1%; p = 0.0078) mutations were significantly depleted in treated patients. APC mutation frequencies were high in both groups and did not differ significantly.

#### Comparison between early– and late-onset Non-Hispanic White patients treated with FOLFOX

In NHW patients treated with FOLFOX (Table S7), no significant differences in WNT gene alterations were observed by age. APC mutations were the most frequent in both EOCRC (77.6%) and LOCRC (76.2%). Other gene alterations, including AMER1, CTNNB1, RNF43, and TCF7L2, occurred at similar rates, indicating a stable WNT mutational profile across age groups in treated NHW patients.

#### Comparison between early– and late-onset Non-Hispanic White patients not treated with FOLFOX

Among untreated NHW patients (Table S8), several statistically significant differences were identified. APC mutations were significantly more common in EOCRC (81.1%) than in LOCRC (73.2%; p = 0.0100). In contrast, AXIN2 (9.8% vs. 5.3%; p = 0.0271) and RNF43 (14.7% vs. 6.6%; p = 5.66×10⁻⁴) mutations were enriched in LOCRC. These findings suggest age-related shifts in canonical versus non-canonical WNT signaling.

#### Comparison between early-onset Hispanic/Latino and Non-Hispanic White patients treated with FOLFOX

Table S9 compares gene-level WNT alterations between FOLFOX-treated EOCRC patients of H/L and NHW ancestry. No significant differences were detected across the eight genes analyzed. APC, CTNNB1, and RNF43 mutation frequencies were nearly identical, suggesting a shared WNT mutational landscape between ancestries under similar treatment conditions.

#### Comparison between untreated early-onset Hispanic/Latino and Non-Hispanic white patients

In contrast, comparison of untreated EOCRC patients (Table S10) revealed ancestry-associated differences. CTNNB1 mutations were significantly more frequent in H/L (17.3%) than NHW (7.6%; p = 0.0467), as were RNF43 mutations (19.2% vs. 6.6%; p = 0.0060). These findings point to enrichment of non-canonical WNT activation in H/L patients and suggest potential ancestry-specific molecular drivers.

#### Comparison between late-onset Hispanic/Latino and Non-Hispanic White patients treated with FOLFOX

Among FOLFOX-treated LOCRC patients (Table S11), WNT mutation frequencies were generally similar between H/L and NHW groups. APC was the most frequently mutated gene in both (71.4% H/L vs. 76.2% NHW). While RNF43 mutations were more common in H/L patients (12.1% vs. 6.5%), this difference was not statistically significant (p = 0.0778).

#### Comparison between late-onset Hispanic/Latino and Non-Hispanic White patients not treated with FOLFOX

In untreated LOCRC patients (Table S12), no statistically significant ancestry-related differences were observed. However, AXIN2 mutations were more frequent in NHW patients (9.8%) compared to H/L (2.0%; p = 0.0748), suggesting possible ancestry-associated variation in non-canonical WNT signaling that warrants further study.

### 3.5 Mutational Landscape

#### Mutational landscape of WNT pathway in early-onset Hispanic/Latino CRC

To characterize the mutational landscape of WNT signaling in EOCRC among Hispanic/Latino (H/L) patients, we analyzed somatic alteration types and FOLFOX treatment status using an oncoplot representation (Figure 1A) alongside detailed mutation classifications (Table S13). Among the 113 EO H/L CRC samples evaluated, 112 (99.1%) harbored at least one mutation in a WNT pathway gene, highlighting the pervasive involvement of this pathway in tumorigenesis in this population.

**Figure 1.**
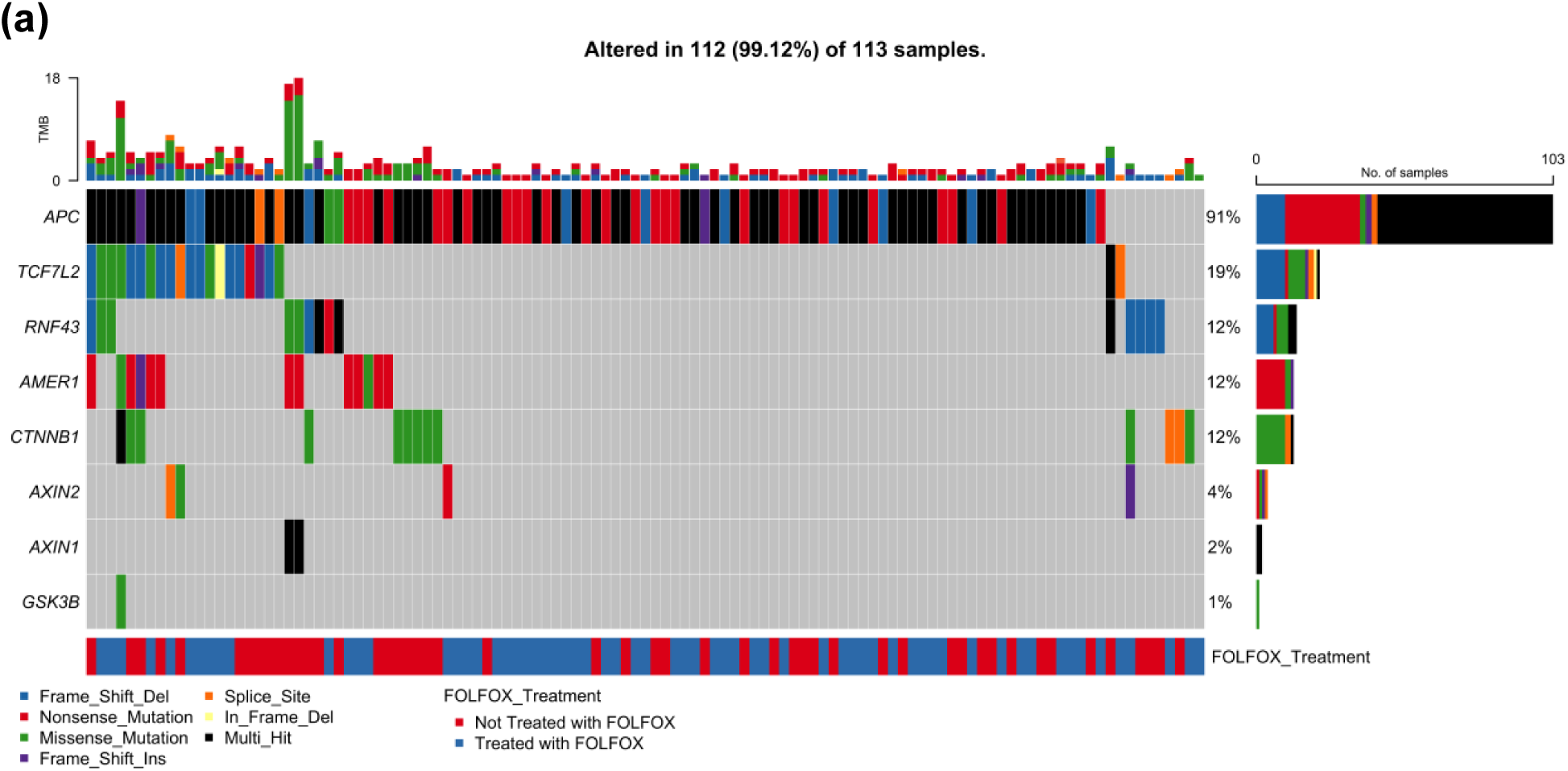
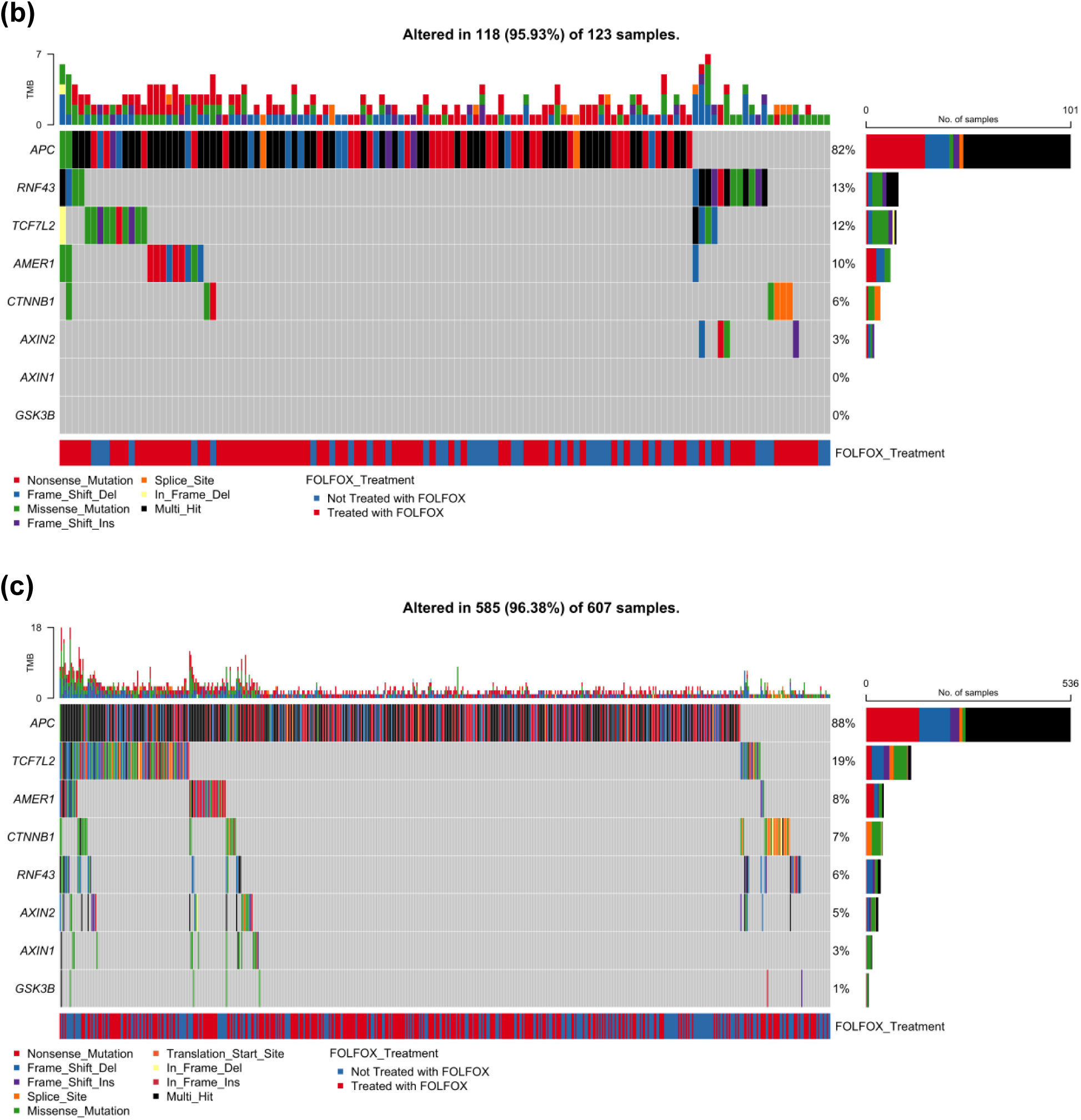
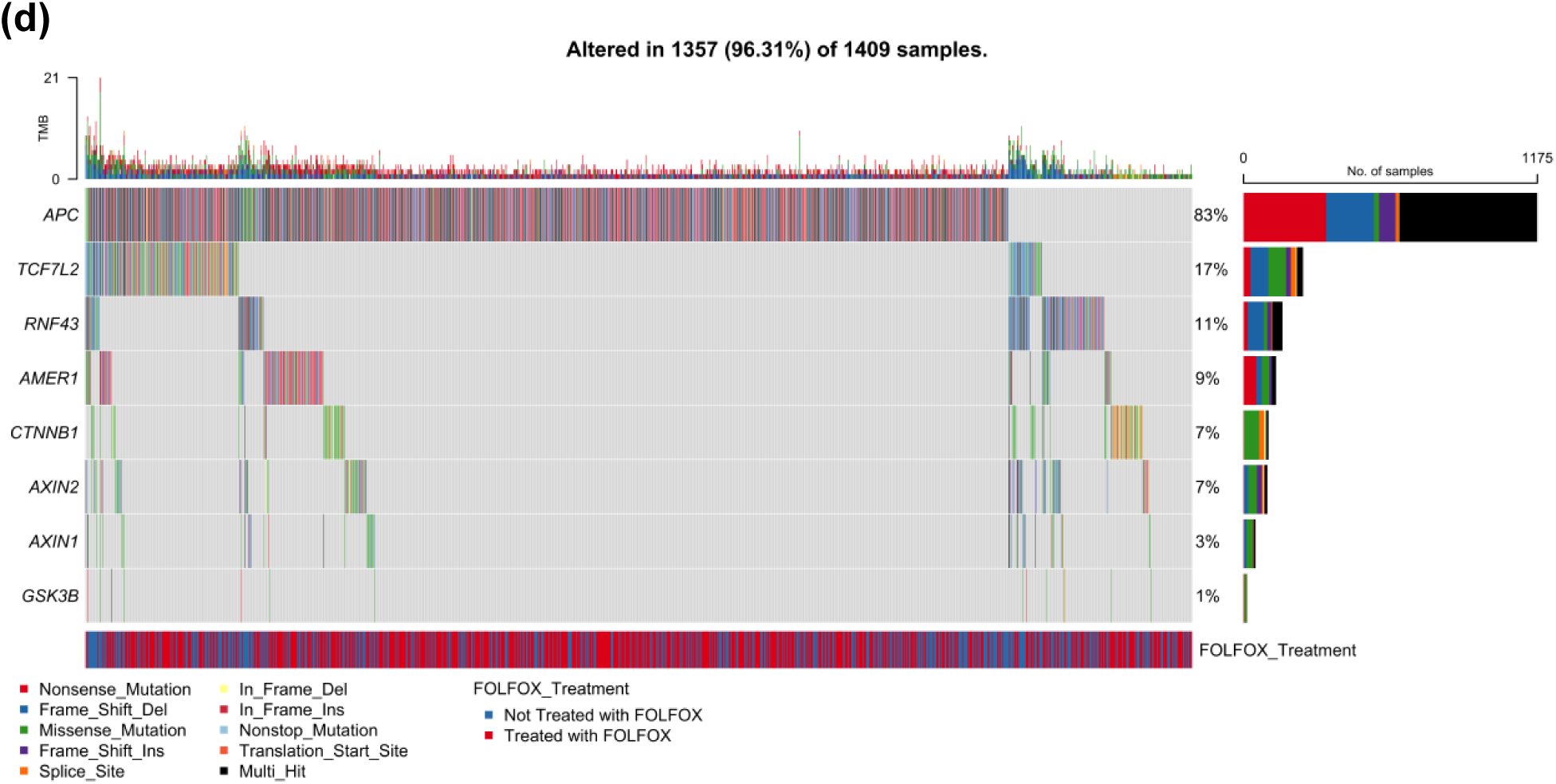
Somatic mutation landscape of WNT pathway genes in colorectal cancer (CRC) stratified by age and ancestry. Oncoplots displaying gene-level mutation profiles of WNT pathway components in colorectal cancer, stratified by age of onset (early vs. late) and ancestry (Hispanic/Latino vs. Non-Hispanic White). Panels show mutation types, tumor mutational burden (TMB), and FOLFOX treatment status across: (a) 113 early-onset Hispanic/Latino (H/L) patients, (b) 123 late-onset H/L patients, (c) 607 early-onset Non-Hispanic White (NHW) patients, and (d) 1,409 late-onset NHW patients. Across all subgroups, *APC* is the most frequently mutated gene, with truncating mutations predominating. Non-canonical WNT genes such as *RNF43*, *TCF7L2*, *AMER1*, and *CTNNB1* exhibit additional recurrent alterations. The data highlight the pervasive disruption of WNT signaling in CRC and suggest age– and ancestry-associated variation in the somatic mutation landscape.

The most frequently mutated gene was APC, altered in 91% of cases. APC mutations were predominantly truncating, including nonsense mutations (52.2%), frame shift deletions (26.1%), and multi-hit events, consistent with its established role as a gatekeeper of WNT signaling. RNF43 mutations were present in 12% of samples and were similarly enriched for truncating alterations, with frame shift deletions (53.3%) and nonsense mutations (20.0%) comprising the majority.

Additional commonly mutated genes included TCF7L2 (19%), AMER1 (12%), and CTNNB1 (12%). TCF7L2 exhibited a diverse spectrum of mutation types, including missense, splice site, and frame shift alterations. AMER1 was enriched for nonsense and missense mutations, while CTNNB1 displayed primarily missense and in-frame deletions. Less frequently altered WNT regulators included AXIN2 (4%), AXIN1 (2%), and GSK3B (1%), with a predominance of missense mutations in AXIN1 (85.7%) and AXIN2 (75.0%).

Tumor mutational burden (TMB), shown in the top barplot of Figure 1A, was generally low across the cohort, though a subset of tumors exhibited elevated TMB, potentially suggestive of mismatch repair deficiency or hypermutator phenotypes. The bottom annotation illustrates FOLFOX treatment status, showing no clear segregation of mutation types between treated and untreated patients.

Overall, these findings underscore a high prevalence of WNT pathway alterations in EO H/L CRC, with a dominant pattern of truncating mutations in APC and diverse disruptions across multiple WNT pathway components, supporting their central role in early-onset tumorigenesis within this high-risk population.

#### WNT pathway alterations in late-onset Hispanic/Latino CRC

To assess the mutational landscape of WNT signaling in LOCRC among Hispanic/Latino (H/L) patients, we analyzed mutation types and treatment status using an oncoplot framework (Figure 1B), complemented by detailed classification of variant types (Table S13). Among 123 LO H/L CRC samples, 118 (95.9%) harbored at least one somatic alteration in a WNT pathway gene, reinforcing the persistent involvement of this pathway in colorectal tumorigenesis across the age spectrum.

APC was again the most frequently mutated gene, altered in 82% of samples. The mutation spectrum was dominated by nonsense mutations (49.3%), frame shift deletions (30.4%), and multi-hit events, underscoring its central role as a truncating tumor suppressor. RNF43, mutated in 13% of cases, displayed a more heterogeneous profile, comprising frame shift deletions (43.8%), nonsense mutations (25.0%), and missense mutations (25.0%), indicating both loss-of-function and potentially modulatory effects.

Additional frequently mutated genes included TCF7L2 (12%) and AMER1 (10%). TCF7L2 alterations encompassed a diverse array of missense, splice site, and frame shift mutations, while AMER1 was enriched for nonsense and missense events. CTNNB1 mutations were observed in 6% of samples and were largely composed of missense and in-frame deletions, consistent with potential functional impacts on β-catenin regulation. AXIN2 was altered in 3% of cases, with a shift toward missense mutations (62.5%) but also a modest representation of truncating mutations. AXIN1 and GSK3B did not exhibit detectable mutations in this cohort. The tumor mutational burden (TMB), shown in the top barplot of Figure 1B, was generally low across the cohort, although a subset of samples displayed elevated TMB, possibly reflecting mismatch repair deficiency or hypermutation. The distribution of FOLFOX treatment status (bottom annotation) revealed no distinct clustering or enrichment of mutation types between treated and untreated patients.

Collectively, these results demonstrate that the WNT signaling axis remains a critical driver of tumorigenesis in LO H/L CRC, with APC truncating mutations as the hallmark event, and additional contributions from diverse alterations in both canonical and non-canonical pathway components. The similarities in mutational architecture between EO and LO H/L CRC suggest a shared molecular vulnerability within this population.

#### WNT pathway alterations in Early-Onset Non-Hispanic White CRC

To delineate the mutational architecture of the WNT signaling pathway in EOCRC among non-Hispanic White (NHW) patients, we analyzed mutation types and FOLFOX treatment status using an oncoplot representation (Figure 1C), supported by mutation classification data (Table S13). Among 607 EO NHW CRC samples, 585 (96.4%) harbored at least one somatic alteration in a WNT pathway gene, confirming the centrality of WNT dysregulation in EOCRC tumorigenesis in this population.

APC mutations were the most frequent, found in 88% of cases, with a predominance of nonsense mutations (56.6%), frame shift deletions (28.3%), and multi-hit events, consistent with its role as a truncating tumor suppressor and negative regulator of WNT signaling. TCF7L2, altered in 19% of samples, showed a broad spectrum of mutation types, including missense, splice site, frame shift deletions, and in-frame insertions, suggesting diverse functional consequences.

RNF43 mutations were present in 6% of samples, largely consisting of frame shift deletions (56.3%) with additional nonsense and missense events. Mutations in AMER1 (8%) and CTNNB1 (7%) were also observed, with AMER1 showing a mixture of nonsense and missense alterations, while CTNNB1 exhibited missense mutations and in-frame deletions that may influence β-catenin activity. AXIN2 was mutated in 5% of cases, with a relatively even distribution of missense (30.0%), frame shift deletion (30.0%), and nonsense mutations (30.0%), indicating a broader mutation spectrum compared to H/L EOCRC cases. AXIN1 (3%) and GSK3B (1%) mutations were infrequent and predominantly missense or truncating in nature.

The tumor mutational burden (TMB), displayed in the top panel of Figure 1C, was generally low across the cohort, although a few samples showed elevated TMB suggestive of hypermutator phenotypes. The FOLFOX treatment status, annotated at the bottom, showed a balanced distribution of treated and untreated cases across mutation types, with no evident treatment-associated clustering.

Together, these findings demonstrate a high prevalence of APC truncating mutations and widespread involvement of other WNT regulators in EO NHW CRC. The broader diversity of mutation types, particularly in genes such as AXIN2 and TCF7L2, suggests a more heterogeneous disruption of WNT signaling in this population, reinforcing the pathway’s central role in early-onset colorectal tumorigenesis.

#### WNT pathway alterations in Late-Onset Non-Hispanic White CRC

To characterize WNT signaling disruptions in LOCRC among non-Hispanic White (NHW) patients, we analyzed 1,409 tumor samples using oncoplot visualization (Figure 1D) and mutation classification data (Table S13). Of these, 1,357 (96.3%) harbored at least one somatic alteration in a WNT pathway gene, reinforcing the pathway’s critical role in LOCRC tumorigenesis in this population.

APC emerged as the most frequently altered gene, mutated in 83% of samples. The majority of APC alterations were truncating in nature, including nonsense mutations (59.5%), frame shift deletions (25.7%), and multi-hit events, consistent with biallelic inactivation of this key tumor suppressor. This mutational pattern mirrored those seen in both younger NHW and Hispanic/Latino (H/L) patients, underscoring APC’s conserved role across demographic groups.

Other WNT pathway components also demonstrated recurrent alterations. TCF7L2 (17%) displayed a diverse mutation spectrum, including missense, splice site, frame shift, and in-frame insertion/deletion mutations, suggesting various functional disruptions. RNF43 was mutated in 11% of samples, with frame shift deletions (61.5%) as the predominant event, alongside nonsense and missense mutations—indicative of its tumor suppressor role through truncating alterations. AMER1 mutations occurred in 9% of samples and were primarily nonsense and missense types. CTNNB1 and AXIN2were each mutated in 7% of samples; CTNNB1 mutations were mostly missense and in-frame deletions, implicating potential dysregulation of β-catenin activity, while AXIN2 exhibited a broader range of mutation types, including missense, frame shift, and splice site variants. AXIN1 mutations (3%) and GSK3B mutations (1%) were less common, with changes predominantly classified as missense or frame shift events. As with earlier-onset cases, AXIN1 and AXIN2mutations were largely missense in nature (87.5% and 70.0%, respectively), indicating a consistent alteration pattern across both age and ethnicity.

Tumor mutational burden (TMB), illustrated in the top panel of Figure 1D, was generally low to moderate, although a subset of samples exhibited elevated TMB, potentially associated with microsatellite instability or hypermutation phenotypes. FOLFOX treatment status, annotated in the lower panel, showed a relatively even distribution of treated and untreated patients across all mutation types, with no distinct clustering by therapy.

In summary, the WNT pathway remains a prominent target of genetic disruption in LO NHW CRC. APC truncating mutations dominate the landscape, complemented by alterations in multiple WNT regulators. The diversity and consistency of mutation patterns across demographic groups highlight the robustness of WNT pathway dysregulation as a key driver of colorectal carcinogenesis and reinforce its relevance for potential therapeutic intervention.

### 3.6 Survival analysis

We evaluated the association between WNT pathway alterations and overall survival across colorectal cancer subgroups defined by age, ancestry, and FOLFOX treatment status using Kaplan-Meier analysis.

Among Hispanic/Latino (H/L) EOCRC patients treated with FOLFOX, no statistically significant difference in overall survival was observed between those with and without WNT pathway alterations (p = 0.89; Figure 2a). Survival trajectories were nearly identical across the follow-up period, with overlapping confidence intervals extending beyond 100 months. Although a minor dip in survival was noted for WNT-altered cases at intermediate time points, this difference diminished over time. Broad confidence intervals—particularly in the unaltered subgroup—reflect small sample sizes and contribute to uncertainty in the survival estimates.

**Figure 2.**
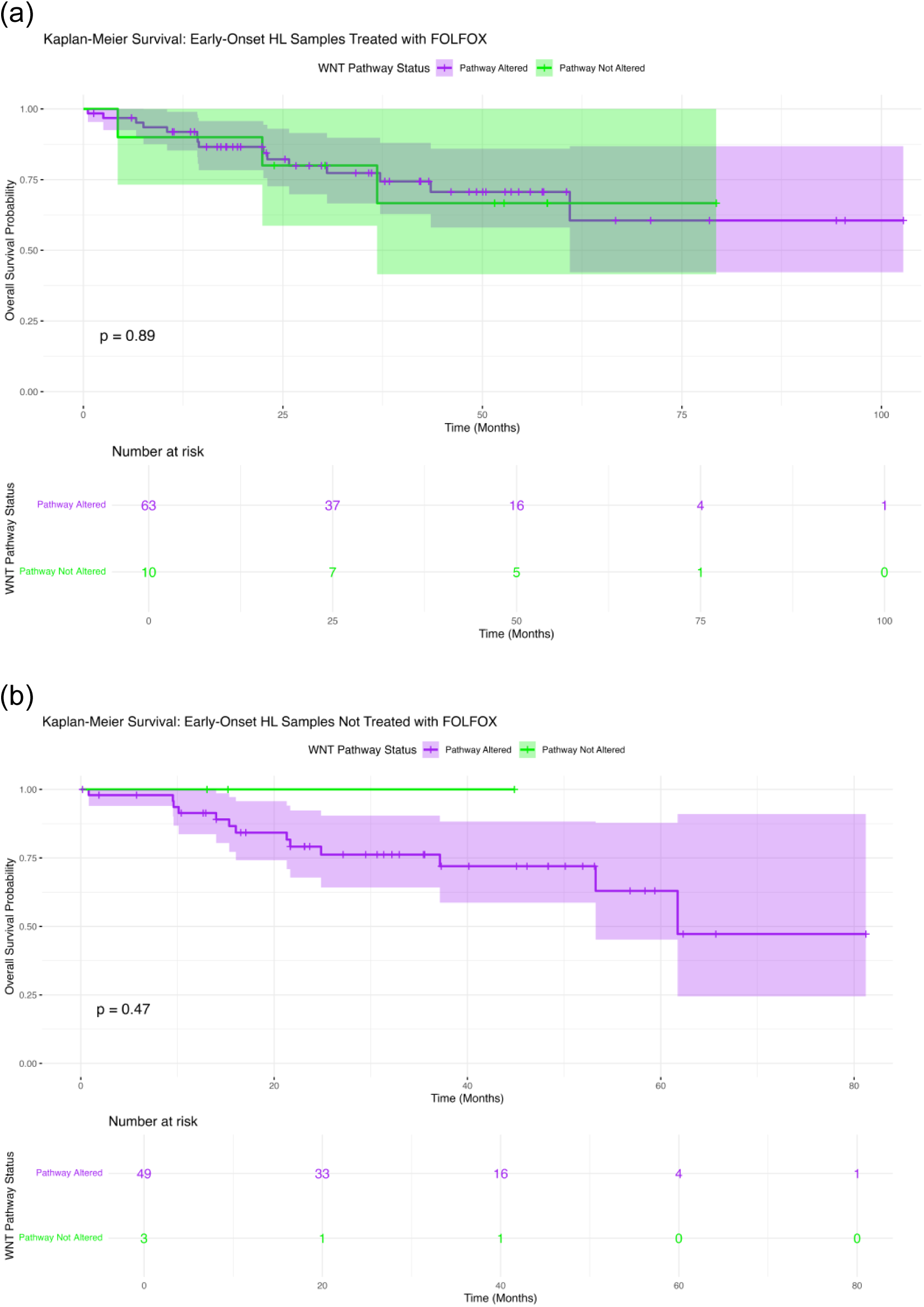
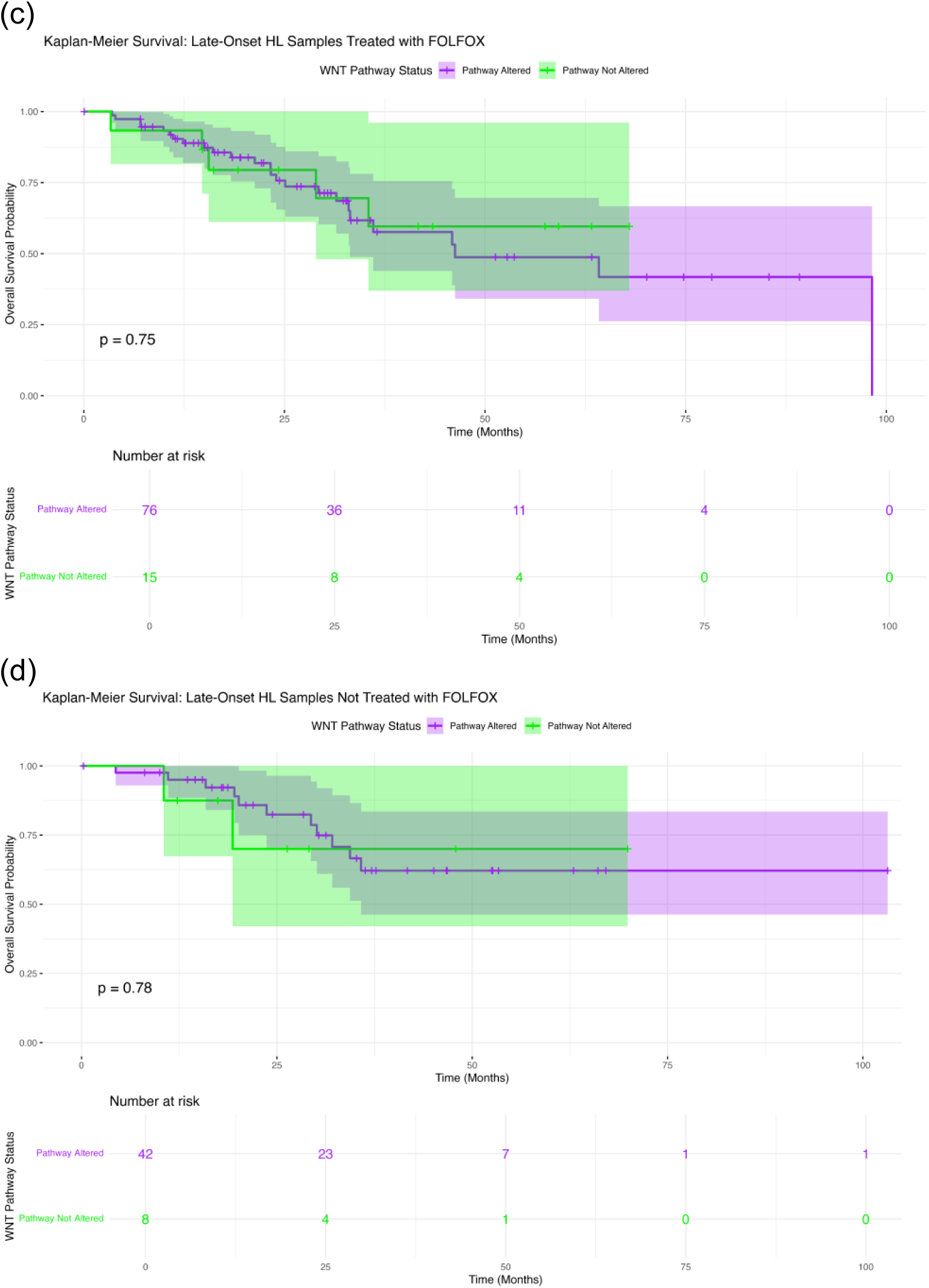
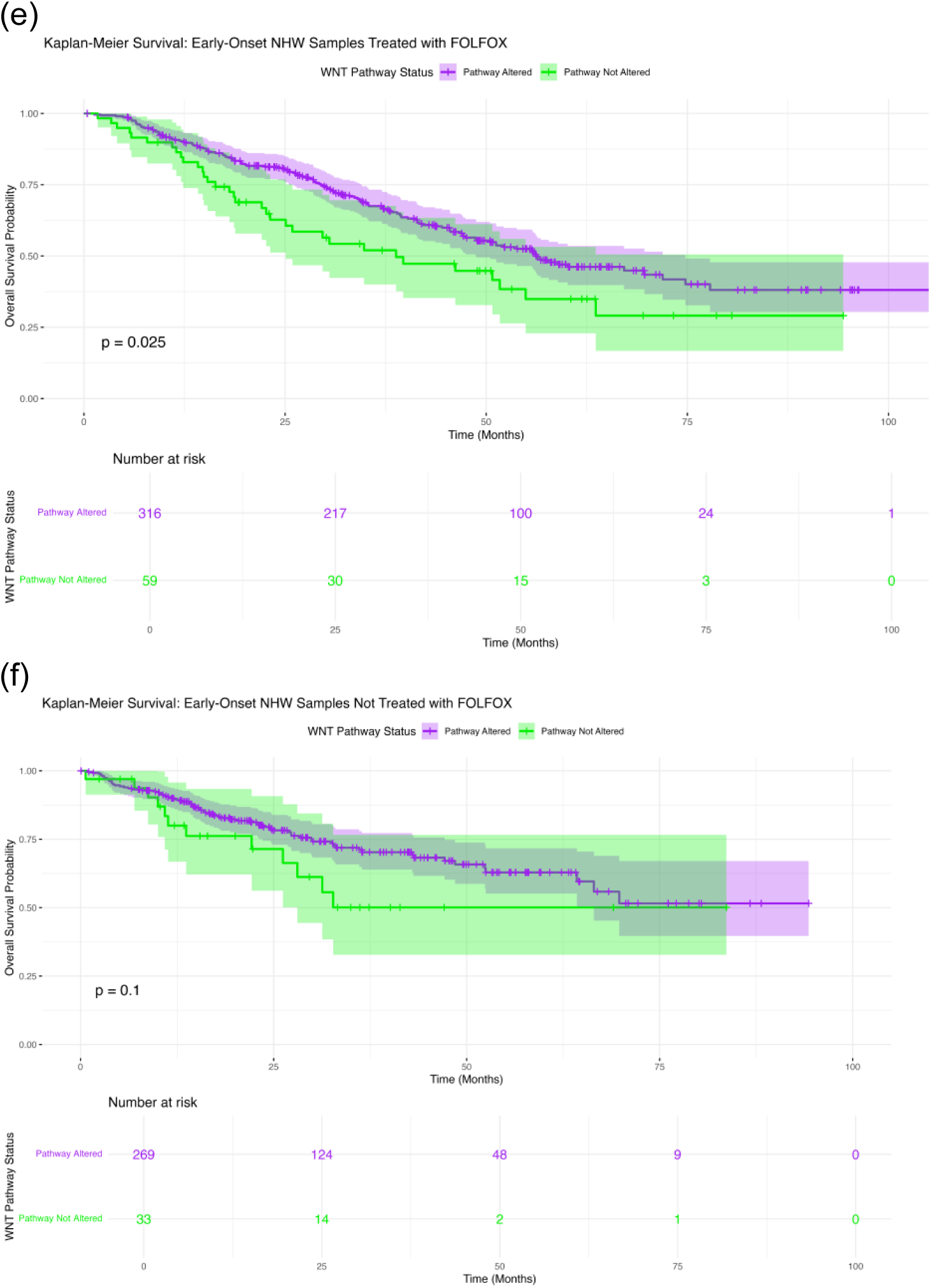
Comparative somatic mutation landscape of WNT pathway genes by age, ethnicity, and FOLFOX treatment status in colorectal cancer. Oncoplots illustrating mutation types and frequencies across WNT pathway genes in six colorectal cancer subgroups: (a) Early-Onset Hispanic/Latino (H/L) Treated with FOLFOX, (b) Early-Onset H/L Not Treated with FOLFOX, (c) Late-Onset H/L Treated with FOLFOX, (d) Late-Onset H/L Not Treated with FOLFOX, (e) Early-Onset Non-Hispanic White (NHW) Treated with FOLFOX, (f) Early-Onset NHW Not Treated with FOLFOX.

Similarly, in H/L EOCRC patients not treated with FOLFOX, WNT alteration status did not significantly impact overall survival (p = 0.47; Figure 2b). Both groups showed comparable survival patterns, with a slight trend toward reduced survival in WNT-altered patients at later follow-up, though this was not statistically significant. Again, wide confidence intervals, especially in the unaltered group, limited interpretability.

For H/L LOCRC patients treated with FOLFOX, survival curves were largely overlapping regardless of WNT mutation status (p = 0.75; Figure 2c). While a modest drop in survival for WNT-altered patients was observed after 75 months, the difference was not statistically significant. Confidence intervals widened notably in later follow-up due to reduced numbers at risk.

In H/L LOCRC patients not treated with FOLFOX, no significant survival differences were detected between WNT-altered and unaltered groups (p = 0.78; Figure 2d). A brief early separation in curves favoring the unaltered group disappeared over time, with substantial widening of confidence intervals beyond 50 months, reflecting small sample sizes and increased uncertainty.

In contrast, among Non-Hispanic White (NHW) EOCRC patients treated with FOLFOX, WNT pathway alterations were associated with significantly improved overall survival (p = 0.025; Figure 2e). Survival curves began diverging early, with WNT-altered patients showing higher survival probabilities that persisted throughout follow-up. Notably, by 50 months, the WNT-unmutated group experienced a sharper decline. Confidence intervals remained relatively narrow until late follow-up, where they widened due to fewer patients at risk.

For NHW EOCRC patients not treated with FOLFOX, no statistically significant survival difference was found (p = 0.1; Figure 2f). However, a trend toward poorer survival was observed in the WNT-unmutated group during the initial follow-up period. This early divergence diminished over time, and broad confidence intervals, particularly in the unaltered group, limited the strength of conclusions.

Among NHW LOCRC patients treated with FOLFOX, WNT pathway alterations were not significantly associated with overall survival (p = 0.42; Figure S1a). Although the WNT-unmutated group showed a slightly lower survival probability early on, curves converged later in follow-up. Confidence intervals widened notably beyond 50 months.

Finally, in NHW LOCRC patients not treated with FOLFOX, WNT pathway alterations were associated with significantly improved overall survival (p = 0.015; Figure S1b). WNT-altered patients maintained higher survival probabilities throughout the follow-up period, with early and persistent separation of survival curves. Confidence intervals remained relatively stable but widened at later time points due to declining numbers at risk.

## 4. Discussion

The rising incidence of early-onset colorectal cancer (EOCRC), particularly in high-risk populations such as Hispanic/Latino (H/L) individuals in Southern California, demands a deeper understanding of the molecular drivers underlying disease onset and treatment response. This study represents one of the largest comparative analyses to date examining the genomic landscape of the WNT signaling pathway in microsatellite stable (MSS) EOCRC, with a specific focus on FOLFOX-treated patients across diverse racial/ethnic groups.

Our findings reveal consistently high frequencies of WNT pathway alterations in EOCRC across both H/L and Non-Hispanic White (NHW) patients, underscoring the centrality of this pathway in early-onset tumorigenesis. APC mutations—primarily truncating—were the most prevalent across all subgroups, reaffirming their role as a foundational driver of WNT dysregulation. However, important differences emerged by age, treatment, and ancestry, suggesting that EOCRC is not a homogeneous disease and that WNT dysregulation may operate via distinct mechanisms in different patient populations.

Among H/L EOCRC patients, FOLFOX-treated tumors exhibited significantly lower frequencies of CTNNB1 and RNF43 mutations compared to untreated cases. This may reflect a selective pressure exerted by chemotherapy or potentially differential sensitivity of specific WNT alterations to FOLFOX-induced cytotoxicity. The reduced prevalence of these mutations following treatment warrants further investigation into whether CTNNB1 and RNF43 alterations confer treatment resistance or are preferentially eliminated by therapy. Conversely, untreated H/L EOCRC patients showed significantly higher frequencies of CTNNB1 and RNF43 mutations compared to their LOCRC counterparts, supporting the hypothesis that EOCRC in this population may be driven by non-canonical WNT activation.

In NHW patients, the mutation landscape also varied by age and treatment status. FOLFOX-treated LOCRC cases demonstrated significantly lower mutation rates in RNF43, AXIN1, AXIN2, and TCF7L2 compared to untreated counterparts, suggesting treatment-related selection or suppression of specific WNT-driven subclones. Interestingly, EOCRC NHW patients exhibited a distinct pattern with significantly lower APC and higher CTNNB1 mutation frequencies relative to LOCRC, further supporting the existence of age-associated divergence in WNT pathway disruption.

Despite the high prevalence of WNT alterations, survival analysis revealed that their prognostic implications were population-specific. In NHW patients, WNT pathway mutations were associated with improved survival in both FOLFOX-treated EOCRC and untreated LOCRC subgroups, suggesting potential value as predictive or prognostic biomarkers. In contrast, WNT alterations did not significantly affect survival outcomes in H/L patients, regardless of treatment or age group. These divergent findings may reflect underlying differences in tumor biology, treatment response, or unmeasured social determinants of health, and underscore the need for population-specific therapeutic strategies.

The diversity of mutation types—particularly in CTNNB1, RNF43, TCF7L2, and AMER1—further highlights the complexity of WNT signaling dysregulation. While APC truncations dominate, non-canonical regulators were variably altered across subgroups, pointing to alternate modes of pathway activation that may influence treatment sensitivity. The consistent prevalence of WNT alterations across treated and untreated patients suggests a foundational role in CRC pathogenesis, but the modulation of specific gene frequencies by FOLFOX raises the possibility of actionable gene-drug interactions.

This study has several limitations. The retrospective design and reliance on publicly available datasets introduce potential biases in clinical annotations and treatment documentation. Additionally, small sample sizes in some subgroups may limit statistical power, particularly for survival analyses in H/L patients. Nonetheless, the use of well-annotated, multi-institutional datasets enabled robust subgroup analyses and highlighted critical differences that merit further validation in prospective studies.

The emergence of next-generation artificial intelligence (AI) agents is poised to transform precision medicine by enabling real-time, integrative analysis of complex clinical and genomic data. In this context, tools like AI-HOPE [28] (Artificial Intelligence for Health Outcomes and Precision Oncology) and AI-HOPE-PM [29] (Artificial Intelligence agent for High-Optimization and Precision mEdicine in Population Metrics) represent a new paradigm in cancer research—facilitating hypothesis generation, pathway interrogation, and biomarker discovery through natural language-driven interfaces. Specifically, AI-HOPE-WNT [30], a specialized agent within this framework, was developed to interrogate WNT signaling alterations in colorectal cancer by integrating multi-omics, treatment, and outcome data. Its application enables rapid stratification of mutation patterns across subgroups such as EOCRC versus LOCRC, and across racial/ethnic populations, thereby accelerating the identification of context-specific therapeutic vulnerabilities. As demonstrated in this study, the complexity and heterogeneity of WNT pathway alterations demand scalable, adaptive tools that can synthesize disparate datasets and inform individualized treatment strategies. The AI-HOPE platform exemplifies how AI-driven agents can complement traditional bioinformatics by providing dynamic, conversational access to precision oncology insights—ushering in an era where AI not only interprets but also interacts with biomedical knowledge to support equitable and evidence-based cancer care.

## 5. Conclusion

In summary, this study demonstrates that WNT pathway alterations are highly prevalent in EOCRC across ancestries and treatment contexts, but their distribution and clinical significance vary by age, ethnicity, and FOLFOX exposure. The differential mutation profiles observed in H/L patients—particularly in CTNNB1 and RNF43—highlight the need for ancestry-informed molecular profiling in EOCRC. Moreover, the association between WNT mutations and improved survival in NHW patients points to their potential utility as predictive biomarkers. These findings support the integration of genomic, demographic, and treatment data to refine precision oncology strategies for EOCRC, especially in underserved and high-risk populations.

## Data Availability

All data used in the present study is publicly available at https://www.cbioportal.org/ and https://genie.cbioportal.org. Additional data can be provided upon reasonable request to the authors.

## Supplementary Materials

**Table S1.** – EO HL Treated with FOLFOX vs EO HL Not Treated with FOLFOX.

| WNT Pathway |  |  |  |
| --- | --- | --- | --- |
| Gene | Early-Onset Hispanic/Latino<br>Treated with FOLFOX<br>n (%) | Early-Onset Hispanic/Latino<br>Not Treated with FOLFOX<br>n (%) | p-value |
| AMER1 Mutation |  |  |  |
| Present | 5 (6.8%) | 8 (15.4%) | 0.2136 |
| Absent | 68 (93.2%) | 44 (84.6%) |  |
| APC Mutation |  |  |  |
| Present | 59 (80.8%) | 44 (84.6%) | 0.756 |
| Absent | 14 (19.2%) | 8 (15.4%) |  |
| AXIN1 Mutation |  |  |  |
| Present | 0 (0.0%) | 2 (3.8%) | 0.416 |
| Absent | 73 (100.0%) | 50 (96.2%) |  |
| AXIN2 Mutation |  |  |  |
| Present | 3 (4.1%) | 1 (1.9%) | 0.6404 |
| Absent | 70 (95.9%) | 51 (98.1%) |  |
| CTNNB1 Mutation |  |  |  |
| Present | 4 (5.5%) | 9 (17.3%) | 0.04052 |
| Absent | 69 (94.5%) | 43 (82.7%) |  |
| GSK3B Mutation |  |  |  |
| Present | 1 (1.4%) | 0 (0.0%) | 1 |
| Absent | 72 (98.6%) | 52 (100.0%) |  |
| RNF43 Mutation |  |  |  |
| Present | 4 (5.5%) | 10 (19.2%) | 0.02157 |
| Absent | 69 (94.5%) | 42 (80.8%) |  |
| TCF7L2 Mutation |  |  |  |
| Present | 11 (15.1%) | 11 (21.2%) | 0.5207 |
| Absent | 62 (84.9%) | 41 (78.8%) |  |

**Table S2.** – LO HL Treated with FOLFOX v LO HL Not Treated with FOLFOX.

| WNT Pathway |  |  |  |
| --- | --- | --- | --- |
| Gene | Late-Onset Hispanic/Latino<br>Treated with FOLFOX<br>n (%) | Late-Onset Hispanic/Latino<br>Not Treated with FOLFOX<br>n (%) | p-value |
| AMER1 Mutation |  |  |  |
| Present | 10 (11.0%) | 2 (4.0%) | 0.2132 |
| Absent | 81 (89.0%) | 48 (96.0%) |  |
| APC Mutation |  |  |  |
| Present | 65 (71.4%) | 36 (72.0%) | 1 |
| Absent | 26 (28.6%) | 14 (28.0%) |  |
| AXIN1 Mutation |  |  |  |
| Present | 0 (0.0%) | 0 (0.0%) | 1 |
| Absent | 91 (100.0%) | 50 (100.0%) |  |
| AXIN2 Mutation |  |  |  |
| Present | 3 (3.3%) | 1 (2.0%) | 1 |
| Absent | 88 (96.7%) | 49 (98.0%) |  |
| CTNNB1 Mutation |  |  |  |
| Present | 5 (5.5%) | 2 (4.0%) | 1 |
| Absent | 86 (94.5%) | 48 (96.0%) |  |
| GSK3B Mutation |  |  |  |
| Present | 0 (0.0%) | 0 (0.0%) | 1 |
| Absent | 91 (100.0%) | 50 (100.0%) |  |
| RNF43 Mutation |  |  |  |
| Present | 11 (12.1%) | 5 (10.0%) | 0.9232 |
| Absent | 80 (87.9%) | 45 (90.0%) |  |
| TCF7L2 Mutation |  |  |  |
| Present | 9 (9.9%) | 6 (12.0%) | 0.9178 |
| Absent | 82 (90.1%) | 44 (88.0%) |  |

**Figure S1.**
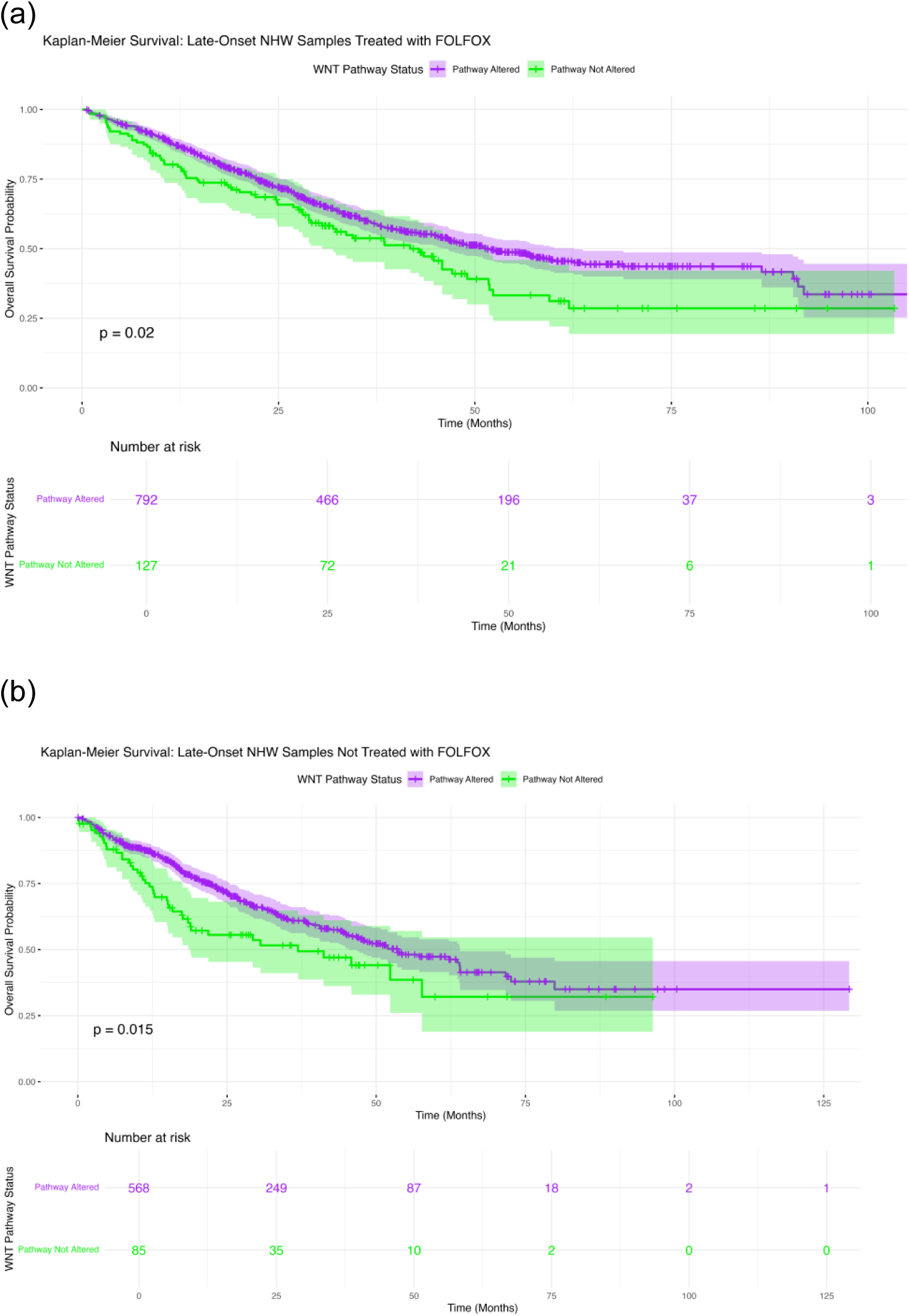
Comparative Somatic Mutation Landscape of WNT Pathway Genes by Age, Ethnicity, and FOLFOX Treatment Status in Colorectal Cancer. Oncoplots illustrating mutation types and frequencies across WNT pathway genes in two colorectal cancer subgroups: (a) Late-Onset NHW Treated with FOLFOX, and (b) Late-Onset NHW Not Treated with FOLFOX.

